# Performance evaluation of novel fluorescent-based lateral immune flow assay (LIFA) for rapid detection and quantitation of total anti-SARS-CoV-2 S-RBD binding antibodies in infected individuals

**DOI:** 10.1101/2022.01.04.22268717

**Authors:** Farah M. Shurrab, Nadin Younes, Duaa W. Al-Sadeq, Hamda Qotba, Laith J. Abu-Raddad, Gheyath K. Nasrallah

## Abstract

1.

**Background:** The vast majority of the commercially available LFIA is used to detect SARS-CoV-2 antibodies qualitatively. Recently, a novel fluorescence-based LIFA test was developed for quantitative measurement of the total binding antibody units (BAU/mL) against the receptor-binding domain of the SARS-CoV-2 spike protein (S-RBD).

**Aim:** To evaluate the performance of the fluorescence LIFA Finecare™ 2019-nCoV S-RBD test along with its reader (Model No.: FS-113).

**Methods:** Plasma from 150 RT-PCR confirmed-positive individuals and 100 pre-pandemic samples were tested by FinCare™ to access sensitivity and specificity. For qualitative and quantitative validation of the FinCar™ measurements, the BAU/mL results of FinCare™ were compared with results of two reference assays: the surrogate virus-neutralizing test (sVNT, GenScript, USA), and the VIDAS®3 automated assay (BioMérieux, France).

**Results:** Finecare™ showed 92% sensitivity and 100% specificity compared to PCR. Cohen’s Kappa statistic denoted moderate and excellent agreement with sVNT and VIDAS®3, ranging from 0.557 (95% CI: 0.32-0.78) to 0.731 (95% CI: 0.51-0.95), respectively. A strong correlation was observed between Finecare™/sVNT (*r=*0.7, p<0.0001) and Finecare™/VIDAS®3 (*r=*0.8, p<0.0001).

**Conclusion:** Finecare™ is a reliable assay and can be used as a surrogate to assess binding and neutralizing antibody response post-infection or vaccination, particularly in none or small laboratory settings.

## 2. Introduction

Soon after the emergence of the severe acute respiratory syndrome 2 (SARS-CoV-2) in December 2019 and its declaration as a pandemic by the World Health Organization (WHO), the need for accurate, sensitive, and rapid detection of SARS-CoV-2 for the control and prevention of the disease was urgent (Theel et al., 2020). Several commercial COVID-19 test kits were developed in response to this urgent need to detect either nucleic acid or antibodies (Van Walle et al., 2021). Although RT-PCR is used as a gold standard test, it requires specialized conditions, expensive equipment, and qualified personnel for sampling and testing. These limitations pose a challenge in a pandemic setting which requires rapid and reliable tests that can be used to screen populations (Dortet et al., 2021). Thus, serological testing, particularly point□of□care approaches, such as the lateral flow immunoassay (LFIA), provides a rapid, portable, and coast effective method complementary to the RT-PCR.

Conventional LFIA used in clinical settings for diagnosing SARS-CoV-2 infection are done manually within 15 minutes, and the results are interrupted qualitatively by the naked eye. As a result, errors due to manual operation and inadequate visual sensitivity interpretation can occur. Recently, Wondfo Biotech developed Fluorescence-based LIFA to detect total anti-S-RBD binding antibodies (BAU) against SARS-CoV-2. Finecare™ 2019-nCoV RBD Antibody Test is a fluorescence immunoassay that is done semiautomatically along with a small portable device. The combination of fluorescence and LIFA provides higher sensitivity, quantitative detection of antibodies. In addition, the test is easily affordable and accessible in small clinical laboratories, research settings, or point of care testing, including post-infection or vaccination.

To the best of our knowledge, there is only one paper in the literature evaluating the rapid fluorescence□based LFIA immunoassays, which aimed to validate the performance of rapid SARS□CoV□2 IgM and IgG test kits based on fluorescence immunochromatography (Kang et al., 2021). This study aimed to evaluate the performance of a rapid semiautomated fluorescence-based LIFA. This assay is designated to provide qualitative and quantitative measurements of SARS□CoV□2 anti-S-RBD total antibodies. One of the most advantages of this assay over the commercially available LIFA is that results are quantitative and converted to BAU/mL (instead of arbitrary ARU/mL) as recently recommended by the WHO.

## 3. Materials and Methods

### 3.1 Study design and ethical approval

We evaluated the performance of FineCare™ 2019-nCOV Antibody test and its reader (Model No.: FS-113); from here on, abbreviated as FineCare™. The serum samples were collected from volunteer individuals between July 26 and September 9, 2020, as a nationwide survey sub-study in Qatar (Al-Thani et al., 2020, Syed et al., 2021). The project was approved by the Institutional Review Boards at Qatar University (QU-IRB 1492-E/21 and QU-IRB 1469-E/21). These samples were used in previous studies (Al-Jighefee et al., 2021, Yassine et al., 2021, Younes et al., 2021).

### 3.2 Sensitivity and specificity determination

The sensitivity FineCare™ was determined using 150 sera from RT-PCR-confirmed individuals (7 - >21 days). Complete descriptions for these participants are summarized in Table S1, and the methodology for PCR detection methodology is described in the following paper (Corman et al., 2020, Yassine et al., 2021). The specificity was examined using 100 pre-pandemic plasma samples collected before 2019, which were used in previous studies (Nasrallah et al., 2018, Nasrallah et al., 2019, Smatti et al., 2020). The panel included plasma samples seropositive for (a) dengue virus (n=26), (B) parvovirus B19 (n = 8), (d) non-respiratory viruses (n =66)

### 3.3 Serological assays

#### 3.3.1 Finecare™ 2019-nCoV RBD Antibody Test

Finecare™ 2019-nCoV RBD Antibody Test is a fluorescence immunoassay technology, specifically the sandwich immunodetection method. It uses fluorescently labeled SARS-CoV-2 S-RBD proteins to form immune complexes by binding to SARS-CoV-2 S-RBD antibodies present in the specimen. It is used along with the Finecare™ FIA Meters (Model No.: FS-113) for the quantitative detection of total antibodies against SARS-CoV-2 S-RBD (Co.). The test can be done using various specimens, including fingerstick whole blood, venipuncture blood, serum, or plasma. There are two test modes for Finecare™ FIA Meters, standard test mode, and quick test mode. The difference is that samples are incubated inside the cartridge holder of Finecare™ FIA Meters in the standard mode, while the samples are incubated at room temperature in the quick mode. The test was done according to the manufacturer’s instructions. In brief, 20 µL of plasma was added to the provided buffer tube and mixed for 45 seconds. Then, 75 µL were added to the test cartridge, incubated for 15 minutes at RT, and then inserted in the test cartridge holder of Finecare™ FIA Meters holder for measurement on quick mode. Finecare™ FIA Meter displays the test result automatically on the screen. The results are given as relative fluorescence units (RFU, AU/mL). This test has a WHO international standardization factor to convert readings from arbitrary units per milliliter (AU/mL) to binding antibody units per milliliter (BAU/mL), 1 AU/mL = 20 BAU/mL. Readings ≥1 AU/mL or ≥20 BAU/ml indicate positive results.

#### 3.3.2 cPass GeneScript sVNT

The FDA-approved SARS-CoV-2 surrogate virus neutralization test (sVNT) developed by GeneScript is widely used in the literature as a reference assay to measure neutralizing antibodies (Cat. No. L00847-C, GenScript Biotech, NJ, USA) (Ismail et al., 2021, Meyer et al., 2020, Younes et al., 2021). This assay detects antibodies that inhibit the interaction between SARS-CoV-2 S-RBD-HRP fusion protein and ACE2 that is coated in a 96-well plate (Meyer et al., 2020). The assay highly correlates with the conventional pseudovirus neutralization test (pVNT, R2 = 0.84) and demonstrated high specificity (99.9%) and sensitivity (100%) (Tan et al., 2020). The test was done according to the manufacturer’s instructions. An inhibition value of ≥ 30% signal inhibition was considered positive, and < 30% signal inhibition was considered negative. The WHO international standardization factor for this assay was used to convert readings from % inhibition to international units per milliliter (IU/mL) by applying the recently published formula (Zhu et al., 2021).

#### 3.3.3 BioMérieux VIDAS®3

VIDAS®3 SARS-CoV-2 IgG (REF 424114) is a widely used automated CE-*in Vitro* diagnostic (IVD) assay and granted the Emergency Use Authorization (EUA) by FDA in early 2021. The assay principle is based on a two-step enzyme immunoassay combined with an enzyme-linked fluorescent assay (ELFA) detection technique. The test uses a solid phase receptacle (SPR) coated with SARS-CoV-2 S-RBD of the spike protein. The test was done according to the manufactures’ instructions. In brief, the test was calibrated with standard (S1), a positive control (C1), and negative control (C2). Then 100µl of the sample was added to the test strip. The results are generated as relative fluorescence value (RFV), and automatically calculated by the instrument; according to the S1 standard and sample RFV, an index value (*i*) is obtained (where *i*□= □RFV_sample_/RFV_S1_). The test is interpreted as negative when *i*□<□1.00 and positive when *i*□≥□ 1.00 (Renard et al., 2021). All readings were standardized to Binding Antibody Units per milliliters (BAU/mL) by applying the WHO International Standard (20.33 BAU/mL) for the VIDAS®3 SARS-COV-2 IgG.

### 3.4 Statistical analysis

Using the RT-PCR as the reference test, sensitivity, specificity, overall percent agreement, and Cohen’s Kappa coefficient were calculated to assess the Performance of FineCare™. Cohen’s kappa coefficient (κ) measures inter-rater reliability as well as the likelihood that an agreement will occur by chance (Ben-David, 2008, McHugh, 2012). A kappa coefficient value of ≤ 0 indicates no agreement, 0.01–0.20 is a poor agreement, 0.21–0.40 is a fair agreement, 0.41– 0.60 is a moderate agreement, 0.61–0.80 is substantial agreement, and 0.81–1.00 is an almost perfect agreement (McHugh, 2012). Concordance analysis between FineCare™ and sVNT and VIDAS®3 were conducted. The concordance measures included overall positive and negative percent agreement and Cohen’s Kappa statistic. The correlation between FineCare™ and each immunoassay was examined using Pearson’s correlation coefficients (r) with 95% confidence interval (95% CI). A coefficient of 0–0.19 suggests a very weak correlation, 0.2–0.39 a weak correlation, 0.40–0.59 a moderate correlation, 0.6– 0.79 as strong correlation and 0.8–1 as very strong correlation (Swinscow and Campbell, 2002). Receiving operating characteristic (ROC) curve and Youden index were conducted to assess the assay threshold (cut-off indices) and identify optimized one and measure the area under the curve (AUC). The relation between AUC and diagnostic accuracy is direct. The larger the AUC, the more accurate a test can be considered in its overall performance. Statistically, an AUC of <0.5 suggests no discrimination, 0.7-0.8 is considered acceptable, 0.8-0.9 is considered excellent, and >0.9 is considered outstanding. To determine the optimal threshold for optimal sensitivity and specificity for FineCare™, the Youden’s index (J) was calculated using the formula: J= max (sensitivity + specificity) – 1, (Fluss et al., 2005, Unal and medicine, 2017). All statistical analyses were performed using GraphPad Prism version 9.3.0.

## 4. Results

### 4.1 Diagnostic Performance of FineCare™ using RT-PCR as a reference test

The overall diagnostic performance of FineCare™ analyzer compared to RT-PCR is summarized in Table1. The of FineCare™ sensitivity and specificity were 92% (95% CI: 86.44% to 95.80%) and 100% (95% CI: 96.38%-100.00%), respectively. The overall percent agreement with RT-PCR was 96.4% (95% CI: 93.2%-98.1%). The Cohen’s Kappa statistic with RT-PCR denoted an excellent agreement with κ coefficient of 0.881 (95% CI: 0.816-0.946).

### 4.2 Concordance assessment between FineCare™ test, sVNT, and VIDAS®3

The tests’ concordance assessment is summarized in Table 2. The overall percent agreement for sVNT and VIDAS®3 was 92% (95% CI: 86.5-95.4) and 94.4% (95% CI: 87.6-97.6), respectively. The positive percent agreement ranged from 95.1% (88.0-98.1) for FineCare™ vs. VIDAS®3 to 97.7% (93.5-99.2) for FineCare™ vs. sVNT. The negative percent agreement from 50% (95% CI: 29-71.0) for FineCare™ vs. sVNT to 88.9% (95% CI: 56.5-98.0) for FineCare™ vs. VIDAS®3. Cohen’s Kappa statistic denoted moderate to excellent agreement and ranged between 0.557 (95% CI: 0.32-0.78); for FineCare™ /sVNT test combination and 0.731 (95% CI: 0.51-0.95) for FineCare™ /VIDAS®3 test combination. Correlation analysis of the readings obtained from FineCare™, sVNT, and VIDAS®3 is illustrated in Figure 1. Both immunoassays showed strong correlation with FineCare™. Pearson’s correlation coefficients (*r*) ranged from 0.70 for FineCare™ /sVNT to 0.8 for FineCare™ / VIDAS®3.

**Figure 1.**
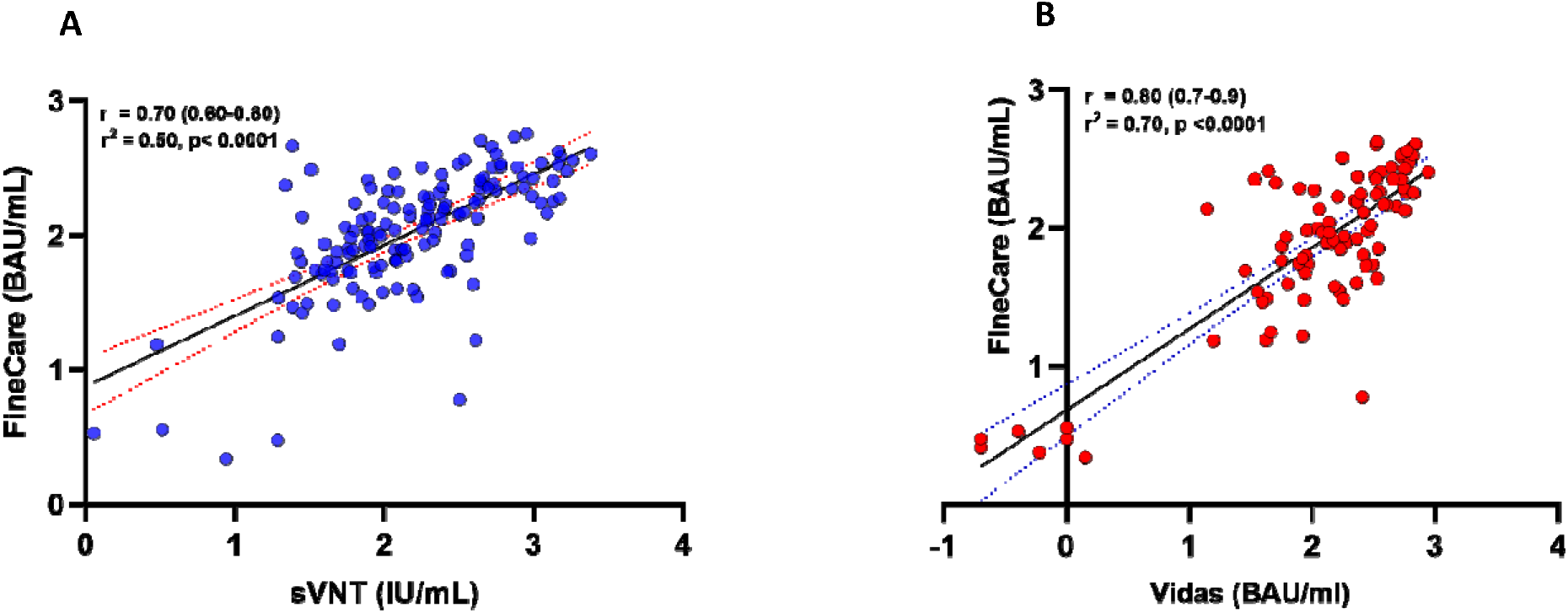
Correlation analysis of the assays standardized readings obtained by each immunoassay. (A) Correlation plot of FineCare™ with the sVNT, (B) Correlation plot of FineCare™ with VIDAS®3. Pearson correlation coefficient (r) and p-value are indicated. Pearson’s r of 0–0.19 is regarded as very weak, 0.2–0.39 as weak, 0.40–0.59 as moderate, 0.6–0.79 as strong and 0.8–1 as very strong correlation, but these are rather arbitrary limits, and the context of the results should be considered. Data are presented for 150 RT-PCR confirmed SARS-CoV-2 positive samples.

### 4.3 Receiver Operating Characteristics (ROC) Curve Analysis

ROC curve analyses showed excellent Performance for FineCare™ with an AUC of 0.9627, and p<0.0001 (Figure 2). Based on the ROC curves and the calculated Youden’s index, the optimized cut-off for detecting total antibodies against RBD was derived. The cut-off obtained was > 10.70, compared to the manufacturer’s cut-off ≥ 20. Applying this new cut-off showed improved sensitivity, from 92% to 94.67%, while the specificity remained 100% (Table 1).

**Table 1:** Diagnostic assessment of FineCare**™** for S-RBD total antibodies detection* RT-PCR used as a reference test

| **Immunoassay** | **FineCare™** |
| --- | --- |
| **Sensitivity % (95% CI)** | 92% (95% CI: 86.44% to 95.80%) |
| **Specificity % (95% CI)** | 100% (95% CI: 96.38%-100.00%) |
| **Overall agreement % (95% CI)** | 96.4% (95% CI: 93.2%- 98.1%) |
| **Cohen's kappa coefficient $\kappa$ (95% CI)** | 0.881 (95% CI: 0.816-0.946) |
| **ROC curves optimized cut-off index** | > 10.70 |
| **Sensitivity using optimized cut-off indices**<br>**% (95% CI)** | 94.67% (95% CI: 89.83%-97.27%) |
| **Specificity using optimized cut-off indices**<br>**% (95% CI)** | 100% (95% CI: 96.34%-100%) |

**Table 2.** Concordance assessment between the FineCare**™** sVNT, VIDAS®3 tests

|  | Compared to | Overall Percent Agreement | Positive Percent Agreement | Negative Percent Agreement | Cohen's kappa coefficient |
| --- | --- | --- | --- | --- | --- |
|  |  | %<br>(95% CI) | %<br>(95% CI) | %<br>(95% CI) | K<br>(95% CI) |
| FineCare™ | sVNT | 92.0%<br>(86.5-95.4) | 97.7%<br>(93.5-99.2) | 50%<br>(29-71.0) | 0.558<br>(0.32-0.78) |
|  | VIDAS®3 | 94.4%<br>(87.6-97.6) | 95.1%<br>(88.0-98.1) | 88.9%<br>(56.5-98.0) | 0.731<br>(0.51 - 0.95) |

**Figure 2.**
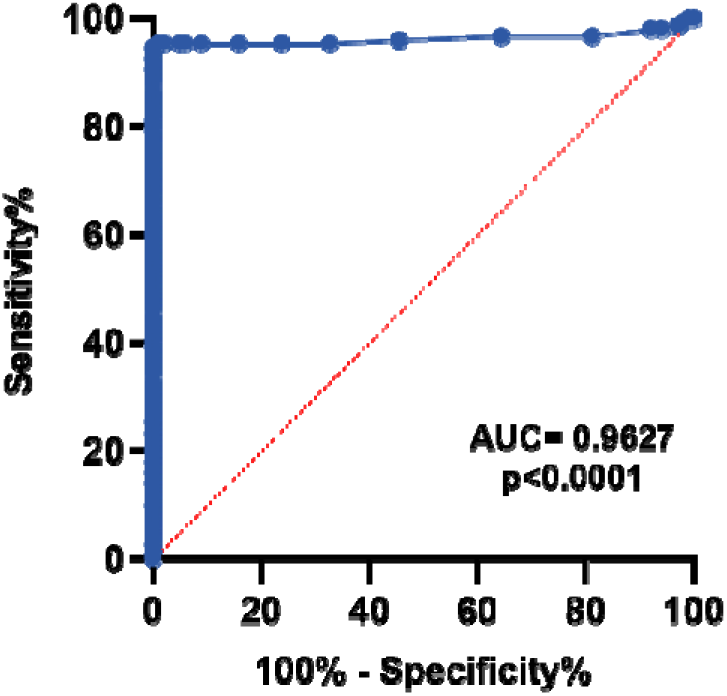
Receiver Operating Characteristic (ROC) curve for Finecare™ 2019-nCoV RBD Antibody Test. An AUC of 0.9–1.0 is considered excellent, 0.8–0.9 very good, 0.7–0.8 good, 0.6–0.7 sufficient, 0.5–0.6 bad, and less than 0.5 considered not useful.

## 5. Discussion

This study validated the performance of Finecare™ 2019-nCoV RBD fluorescence immunoassay for detecting total antibodies against SARS-CoV2 S-RBD after infection. A panel of 150 samples collected from RT–PCR confirmed individuals and 100 pre-pandemic sera were used to evaluate the assays’ performance. To our knowledge, this is the first study conducted to validate the fluorescence-FIA-based FineCare™ 2019-nCoV RBD Antibody test, which marks the novelty of this research work.

Finecare™ test demonstrated high sensitivity and specificity comparable to sVNT and VIDAS®3 and other immunoassays in the market. VIDAS®3 was reported to have a sensitivity and specificity of 88.3% and 98.4%, respectively (Younes et al., 2021). Similarly, sVNT reported a high specificity (99.9%) and sensitivity (95.0–100%) (Tan et al., 2020). It was noticed that out of the 12 negative samples by Finecare™, eight of them were asymptomatic. Earlier studies reported that the severity of the case affects the humoral response in patients (Wu et al., 2021, Zhao et al., 2020). Previously, we reported that the sensitivity of the immunoassay was higher in symptomatic patients than in the asymptomatic patient group (Younes et al., 2021).

We evaluated the degree of correlation between FineCare™ with sVNT and VIDAS®3. A strong correlation was obtained between FineCare™ and both assays (Figure1); however, VIDAS®3 showed a slightly higher correlation. This is because both VIDAS®3 and FineCare™ detection methods are based on enzyme-linked fluorescent and target antibodies against the S-RBD domain. This is particularly important; the FineCare™ rapid antibody test could detect neutralizing antibodies and serve as a surrogate for the advanced automated immunoassays in clinical settings to measure the humoral immune response after vaccination or infection and research context. ROC curve analysis was also performed to determine the optimal cut-off indices for FineCare™ (Figure 2). The new cut-off value showed improved sensitivity without affecting the specificity. However, the cut-off values could be adjusted depending on the clinical setting or research context. For instance, in high-prevalence settings, higher thresholds may be desirable for screening purposes, whereas lower cut-off may be helpful for diagnosis purposes (Ismail et al., 2021).

Our study had some limitations; most of our RT-PCR samples were collected from asymptomatic individuals (Table S1), which might have underestimated the assay’s sensitivity. In addition, the control group did not include samples for other coronaviruses or influenza that might cross-react with SARS-CoV-2, which could have led to an overestimated specificity.

In conclusion, our data showed that FineCare™ 2019-RBD antibody test demonstrated excellent performance in terms of sensitivity, specificity, and overall agreement with RT-PCR as a reference test. In addition, correlation with the FDA-approved sVNT from Genscript and the automated analyzer VIDAS®3 from bioMérieux, FineCare™ immunoassay showed an outstanding performance in detecting total antibodies in serum samples against SARS-CoV-2. Thus, this assay could be reliable for the quantitative detection of antibodies in the vaccinated population and recovered patients.

## Data Availability

All data produced in the present study are available upon reasonable request to the authors

## Author contributions

Conceptualization: GKN. Participant recruitment and demographic data collection: HQ. Laboratory testing: FMS, DWA, NY. Supervision: GKN. Data analysis: GKN, FMS, NY. First draft writing: GKN, FMS. Review and editing: LJA, GKN. Funding: GKN, LJA

## Acknowledgment

we would like to thank Qatar National Research Fund (a member of Qatar Foundation) for funding this work.

## Ethical statement

The study was conducted according to the guidelines of the Declaration of Helsinki and was reviewed and approved by the Institutional Review Board at Qatar University (QU-IRB 1492-E/21 and QU-IRB 1469-E/21). Informed consent was obtained from all subjects involved in the study before collecting samples.

## Data avalibility

All data produced in the present study are available upon reasonable request to the authors

## References

1. 2019-nCoV RBD Antibody Tes; Available from: https://corona-rapid-testing.at/wp-content/uploads/2021/04/finecare-2019-ncov-rbd-antibody-test-instructions.pdf. [Accessed 27/12/2021 2021].

2. 2019-nCoV RBD Antibody Test; Available from: www.wondfo.com.cn. [Accessed 12/26/2021 2021].

3. Al-Jighefee HT, Yassine HM, Nasrallah GKJP. Evaluation of antibody response in symptomatic and asymptomatic COVID-19 patients and diagnostic assessment of new IgM/IgG ELISA kits. 2021;10(2):161.

4. Al-Thani MH, Farag E, Bertollini R, Al Romaihi HE, Abdeen S, Abdelkarim A, et al. Seroprevalence of SARS-CoV-2 infection in the craft and manual worker population of Qatar. 2020.

5. Ben-David AJESwA. Comparison of classification accuracy using Cohen’s Weighted Kappa. 2008;34(2):825–32.

6. Co. WB. Available from: https://en.wondfo.com.cn/pt/index17.html. [Accessed 1/3/2022 2022].

7. Corman VM, Landt O, Kaiser M, Molenkamp R, Meijer A, Chu DK, et al. Detection of 2019 novel coronavirus (2019-nCoV) by real-time RT-PCR. 2020;25(3):2000045.

8. Dortet L, Ronat J-B, Vauloup-Fellous C, Langendorf C, Mendels D-A, Emeraud C, et al. Evaluating 10 commercially available SARS-CoV-2 rapid serological tests by use of the STARD (Standards for Reporting of Diagnostic Accuracy Studies) method. 2021;59(2):e02342–20.

9. Fluss R, Faraggi D, Reiser Bjbjjommib. Estimation of the Youden Index and its associated cut-off point. 2005;47(4):458–72.

10. Ismail A, Shurrab FM, Al-Jighefee HT, Al-Sadeq DW, Qotba H, Al-Shaar IA, et al. Can commercial automated immunoassays be utilized to predict neutralizing antibodies after SARS-CoV-2 infection? A comparative study between three different assays. 2021.

11. Kang K, Huang L, Ouyang C, Du J, Yang B, Chi Y, et al. Development, performance evaluation, and clinical application of a Rapid SARSLCoVL2 IgM and IgG Test Kit based on automated fluorescence immunoassay. 2021;93(5):2838–47.

12. McHugh MLJBm. Interrater reliability: the kappa statistic. 2012;22(3):276–82.

13. Meyer B, Reimerink J, Torriani G, Brouwer F, Godeke G-J, Yerly S, et al. Validation and clinical evaluation of a SARS-CoV-2 surrogate virus neutralisation test (sVNT). 2020;9(1):2394–403.

14. Nasrallah GK, Dargham SR, Mohammed LI, AbuLRaddad LJJJomv. Estimating seroprevalence of herpes simplex virus type 1 among different Middle East and North African male populations residing in Qatar. 2018;90(1):184–90.

15. Nasrallah GK, Dargham SR, Sahara AS, Elsidiq MS, Abu-Raddad LJJJoCV. Performance of four diagnostic assays for detecting herpes simplex virus type 2 antibodies in the Middle East and North Africa. 2019;111:33–8.

16. Renard N, Daniel S, Cayet N, Pecquet M, Raymond F, Pons S, et al. Performance Characteristics of the Vidas SARS-CoV-2 IgM and IgG Serological Assays. 2021;59(4):e02292–20.

17. Smatti MK, Nasrallah GK, Al Thani AA, Yassine Hmjv. Measuring influenza hemagglutinin (HA) stem-specific antibody-dependent cellular cytotoxicity (ADCC) in human sera using novel stabilized stem nanoparticle probes. 2020;38(4):815–21.

18. Swinscow TDV, Campbell MJ. Statistics at square one: Bmj London, 2002.

19. Syed MA, Al Nuaimi AS, Nasrallah GK, Althani AA, Yassine HM, Zainel AA, et al. Epidemiology of SARS-CoV2 in Qatar’s primary care population aged 10 years and above. 2021;21(1):1–11.

20. Tan CW, Chia WN, Qin X, Liu P, Chen MI-C, Tiu C, et al. A SARS-CoV-2 surrogate virus neutralization test based on antibody-mediated blockage of ACE2–spike protein–protein interaction. 2020;38(9):1073–8.

21. Theel ES, Slev P, Wheeler S, Couturier MR, Wong SJ, Kadkhoda KJJocm. The role of antibody testing for SARS-CoV-2: is there one? 2020;58(8):e00797–20.

22. Unal IJC, medicine mmi. Defining an optimal cut-point value in ROC analysis: an alternative approach. 2017;2017.

23. Van Walle I, Leitmeyer K, Broberg EKJE. Meta-analysis of the clinical performance of commercial SARS-CoV-2 nucleic acid and antibody tests up to 22 August 2020. 2021;26(45):2001675.

24. Wu J, Liang B, Chen C, Wang H, Fang Y, Shen S, et al. SARS-CoV-2 infection induces sustained humoral immune responses in convalescent patients following symptomatic COVID-19. 2021;12(1):1–9.

25. Yassine HM, Al-Jighefee H, Al-Sadeq DW, Dargham SR, Younes SN, Shurrab F, et al. Performance evaluation of five ELISA kits for detecting anti-SARS-COV-2 IgG antibodies. 2021;102:181–7.

26. Younes S, Al-Jighefee H, Shurrab F, Al-Sadeq DW, Younes N, Dargham SR, et al. Diagnostic efficiency of three fully automated serology assays and their correlation with a novel surrogate virus neutralization test in symptomatic and asymptomatic SARS-COV-2 individuals. 2021;9(2):245.

27. Zhao J, Yuan Q, Wang H, Liu W, Liao X, Su Y, et al. Antibody responses to SARS-CoV-2 in patients with novel coronavirus disease 2019. 2020;71(16):2027–34.

28. Zhu F, Althaus T, Tan CW, Costantini A, Chia WN, Chau NVV, et al. WHO international standard for SARS-CoV-2 antibodies to determine markers of protection. 2021.

